# Shared Strides: Community-based, high-throughput biomechanics data collection in knee osteoarthritis

**DOI:** 10.64898/2026.03.23.26349064

**Authors:** Jenna M. Qualter, Ryan C. McCloskey, Kathryn A. Stofer, Peihua Qiu, Zibo Tian, Heather K. Vincent, Kerry E. Costello

## Abstract

**Objective:** This analysis assessed the acceptability and recruitment implications of a high-throughput, community-based biomechanics protocol among individuals with knee osteoarthritis (OA).

**Design:** During the Shared Strides Study, high-throughput markerless biomechanics assessment was conducted at community sites to help facilitate research engagement in the OA population. In this cross-sectional study, biomechanics data during a set of activities of daily living (ADLs) and questionnaire data were collected. Adults aged 40 years or older with knee OA participated at one of four sites across Gainesville, FL—two on-campus and two community-based. Eligible individuals were either screened over the phone and scheduled for a specific date and time or screened on site for potential same-day participation. Participant acceptability of the community-based biomechanics data collection approach was assessed using a 15-item custom questionnaire. Recruitment characteristics and participant preferences were compared across sites.

**Results:** The high-throughput community-based data collection approach was well received. Compared with on-campus sites, community-based sites had higher engagement from walk-in participants and new research participants (40% of the sample). Familiarity with, and distance to, a data collection site were important factors in research engagement in this population. No differences in demographic characteristics existed between sites (p > 0.05), but recruitment resulted in a large sample size (n = 85) likely representative of the communities surrounding the selected sites.

**Conclusions:** Integrating markerless motion capture with a community-based research approach may enhance the participant experience and facilitate larger, more heterogeneous sample sizes, ultimately reducing bias and homogeneity in current OA biomechanics research.

## INTRODUCTION

Mechanical loading is one of the few modifiable risk factors implicated in the progression of knee osteoarthritis (OA);^1^ however, patient responsiveness to biomechanical interventions targeting joint loading has been inconsistent,^2^ likely reflecting the substantial inter-individual variability in symptoms, structural damage, and functional limitations among individuals with knee OA.^3^ Historically, this variability has been difficult to account for in biomechanics research due to data collection constraints that result in small sample sizes (typically 2-20 participants) and influence population composition.^4^ To further advance OA biomechanics, large and representative datasets are needed to evaluate biomechanics in the context of known patient heterogeneity.

Traditional laboratory-based, optical motion capture studies are time consuming and typically require participants to travel to laboratory locations for extended data collection sessions. As a result, these studies are often prone to selection bias and yield relatively small and homogeneous samples, limiting the generalizability of results.^5^ In contrast, the development and validation of markerless motion capture^6, 7^ enables more efficient data collection without the need for markers, wearable sensors, or constrictive clothing,^8, 9^ and its potential for large-scale biomechanics data collection outside of traditional laboratory settings has been demonstrated in a general population for walking tasks.^10^ Conducting research in more familiar and accessible locations may reduce barriers to participation associated with traditional laboratory settings.^11, 12^ Although this approach has not been systematically studied in biomechanics research, community-based research in other contexts has demonstrated increased participation among older adults^13^ and groups traditionally underrepresented in research,^14, 15^ suggesting it may facilitate recruitment of more representative samples of individuals with OA to biomechanics studies.

Therefore, the objectives of this study were to: 1) compare recruitment outcomes and characteristics of participants recruited to a markerless biomechanics study across four distinct on-campus and community sites, and 2) assess the acceptability of this high-throughput, community-based biomechanics data collection protocol among individuals with knee OA.

## METHODS

### Study Overview

The current analysis is part of the Shared Strides Study, a feasibility study evaluating high-throughput biomechanics data collection at community sites in individuals with knee OA. In Shared Strides, a portable biomechanics laboratory was transported to and deployed at multiple on-campus and community sites for single-day data collection “events” on weekdays between the hours of 8AM and 5PM. This system consisted of markerless motion capture equipment/software (Vicon Vero camera Nexus 2.16 software, Vicon Motion Systems, Oxford, UK; FLIR cameras, Teledyne FLIR LLC, Arlington, VA, USA; Theia 3D software, Theia Markerless Inc., Kingston, ON, Canada), force platforms embedded in a portable walkway (AMTI, Watertown, MA, USA), and a custom-built mobile computing workstation. Participants were enrolled through a combination of scheduled appointments and on-site walk-in screening during data collection events. Biomechanics and survey-based data were collected from multiple participants per day, with arrivals spaced 15-30 minutes apart. Average visit length was 65.8 ± 16.0 minutes per person (full timing details reported elsewhere), thus multiple participants could be present on site at any given time.

Participants completed an initial visit and a 6-month follow-up visit; the current analysis includes data from the initial visit only. All procedures were approved by the University of Florida Institutional Review Board (#IRB202400674) in accordance with the ethical standards of the responsible committee on human experimentation (institutional and national) and with the Helsinki Declaration of 1975, as revised in 2000. All participants provided written informed consent prior to participation in data collection procedures and received monetary compensation of 20 dollars per visit completed.

### Data Collection Sites

Four data collection sites across the University of Florida campus and the surrounding community in Gainesville, FL were utilized for the Shared Strides Study, including two on-campus and two off-campus locations (Figure 1). Room visibility, parking options, and accessibility varied by site (Table 1). At the community sites, event rooms were visible to foot traffic in adjacent public hallways and activity spaces, creating opportunities for walk-in participation. In contrast, the on-campus sites offered greater privacy: the Clinical Laboratory site was located off a private hallway behind a clinic check-in area, and the Student Union site experienced minimal foot traffic from the target population.

**Figure 1.**
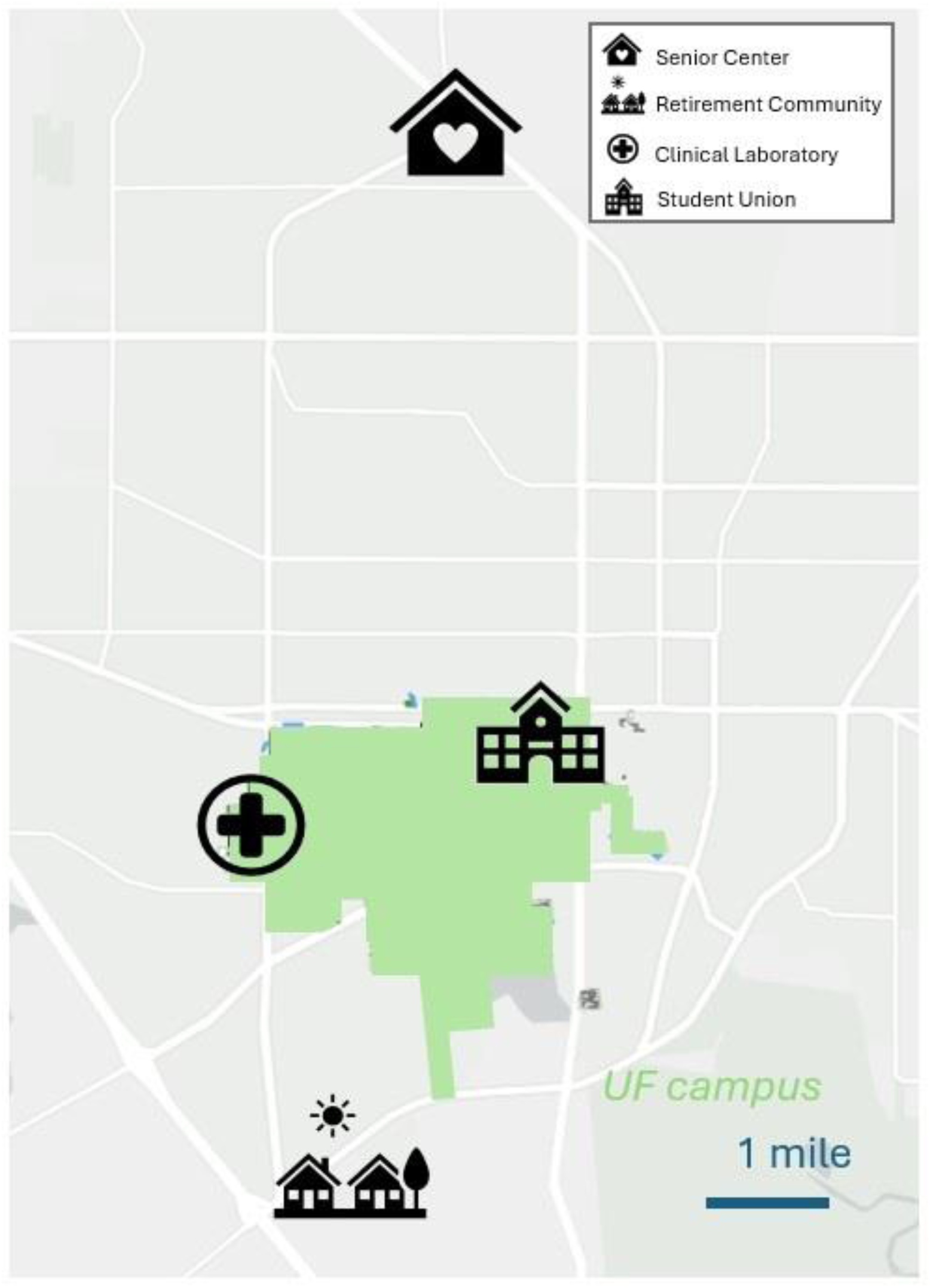
Map of Shared Strides Study site locations across Gainesville, Florida relative to the university campus (shown in green overlay).

**Table 1.**
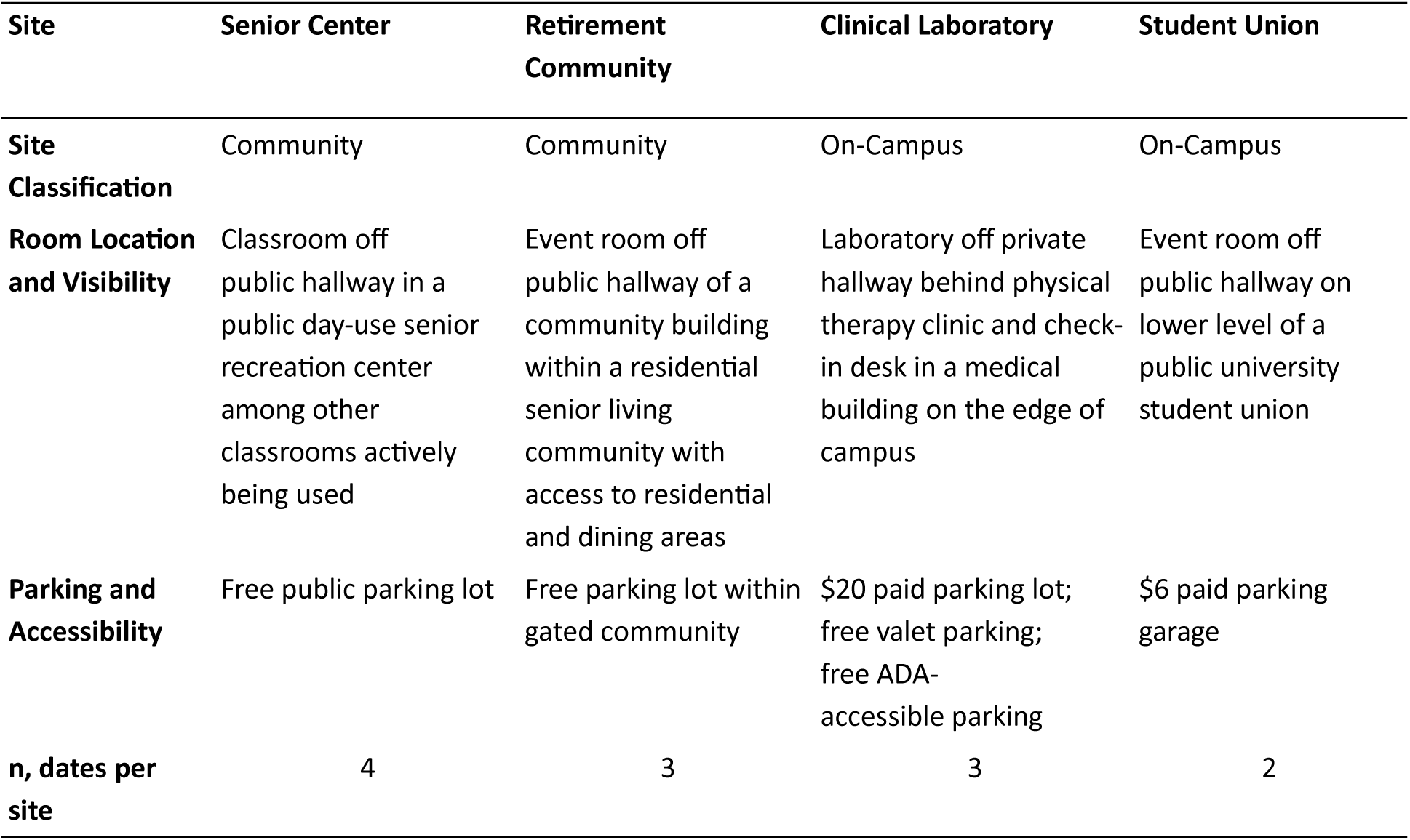
Site characteristics and descriptions.

### Recruitment and Screening

Recruitment occurred over an 11-month period using multiple standard approaches. Study flyers were distributed at community sites and posted throughout the local area. Postcards were mailed to potentially eligible individuals enrolled in a local research registry, and online social media advertisements were used to target individuals in the surrounding region. During data collection events, flyers were also displayed within the data collection sites, allowing interested individuals to inquire about participation on site. Recruitment materials are available in the online supplement (APPENDIX A-B).

Eligible participants were either pre-screened via phone and scheduled for a specific site and date/time based on preference or screened on site as walk-in participants. Based on availability, those screened on site were given the option to participate immediately following screening, to return at a designated time on the same day, or to schedule a future appointment. Additional dates were scheduled at specific locations based on demand.

Individuals aged 40 years or older were considered eligible for the Shared Strides Study if they: (1) met the American College of Rheumatology (ACR) criteria^16^ or (2) self-reported a prior physician diagnosis of OA in one or both knees. Participants were required to be able to walk 20 feet without assistive devices. Participants were excluded if they self-reported preexisting neurological or musculoskeletal conditions (e.g., stroke, lower-back pain) that affected safety and/or impacted movement more than the presence of knee OA, if they had sustained a lower-limb injury or undergone surgery within the 3 months prior to screening, or if they were pregnant.

### Data collection procedures

Data collection procedures were designed to support high-throughput assessment while accommodating variability in participant arrival times and site-specific logistics. Upon arrival at the site, participants completed the informed consent process with a member of the study staff. Following consent, height and weight were recorded and used to calculate body mass index (BMI). The order of the remaining segments (i.e., biomechanics and questionnaires) varied based on scheduling and equipment availability (Figure 2).

**Figure 2.**
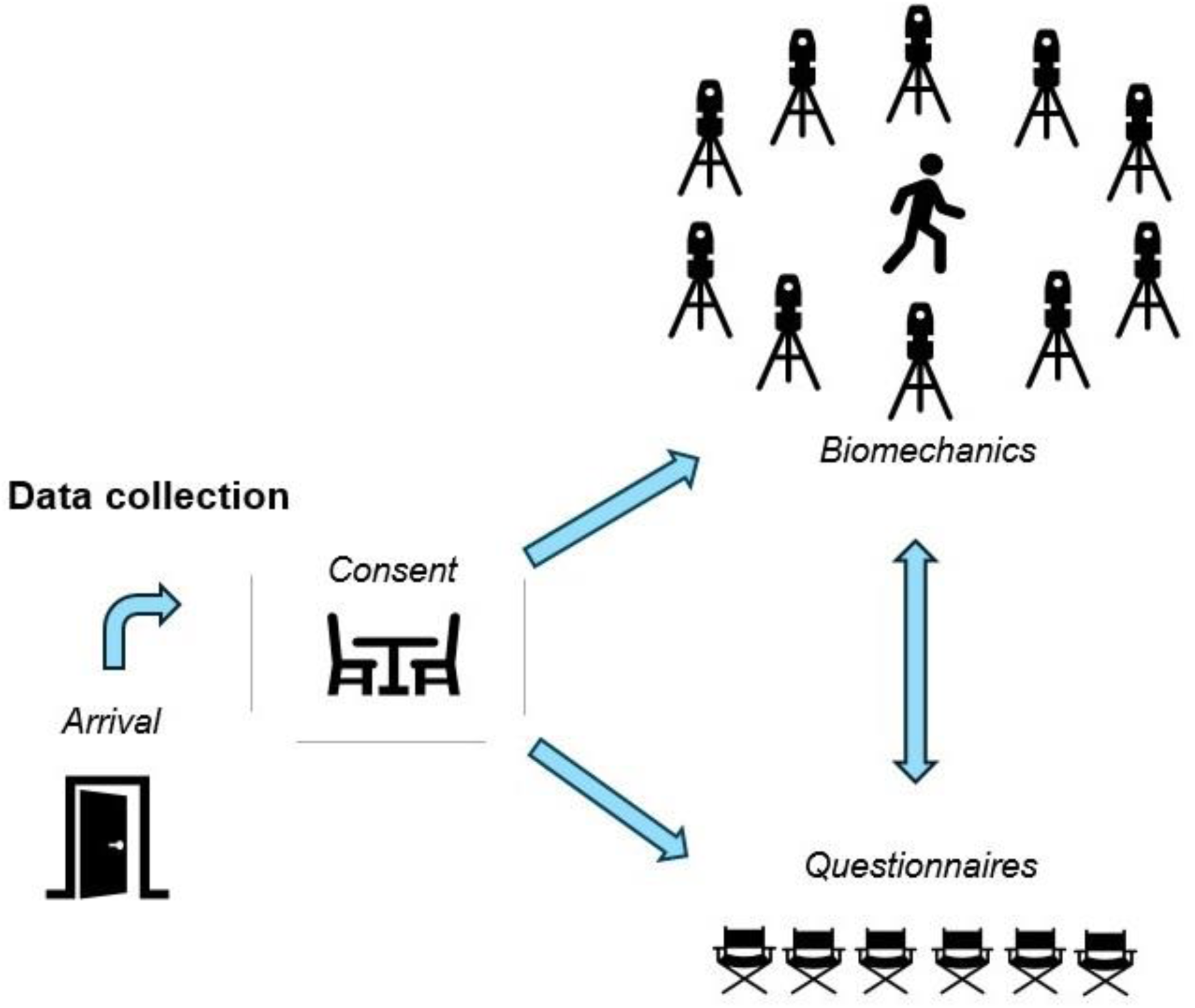
Data collection overview.

During the biomechanics testing, participants performed a set of activities of daily living, including walking at a self-selected usual pace and a self-selected fastest safe pace, double- and single-leg standing balance, squatting, sit-to-stand, step-up, and step-down tasks. Multiple attempts were permitted to obtain three successful trials for each activity, and activities could be skipped at the discretion of the research team (largely due to balance concerns) or at the participant’s request.

Acceptability measures and data collection preferences were assessed with a custom questionnaire designed to evaluate participant perceptions of the high-throughput, community-based biomechanics data collection process (APPENDIX C). This exploratory questionnaire has not been formally validated and may include some ambiguity in interpretation. The questionnaire collected information on prior research participation, transportation to the data collection site (travel time and mode of transportation), likelihood of participating at community versus on-campus sites and reasons for site preference (e.g., familiarity, distance, and transportation/parking options), and motivations for research participation (e.g., altruism, personal benefit, increased knowledge, and compensation). Participants were also asked whether choices in location and date influenced their desire or ability to participate.

Following completion of the acceptability questionnaire, participants provided demographic information and completed a custom clinical history questionnaire focused on knee joint health. OA symptom severity was assessed in the OA knee (left, right, or both) using the Knee Injury and Osteoarthritis Outcome Score (KOOS)^17, 18^, and further patient-reported outcome measures were collected using additional validated questionnaires (e.g., Measure of Intermittent Constant Osteoarthritis Pain^19^). In total, questionnaires consisted of 6 instruments with 100 items for those with unilateral OA and 153 items for those with bilateral OA. All questionnaires were completed electronically on study iPads using REDCap (Research Electronic Data Capture).

### Statistical Analysis

Analyses were designed to descriptively compare recruitment outcomes and acceptability measures across sites in this exploratory, hypothesis-generating feasibility study. Participant characteristics (demographics, recruitment, and acceptability) were compared across the four distinct sites using Welch’s ANOVA (continuous variables) and Fisher’s exact test (categorical variables) to account for the small and unbalanced sample sizes. Assumptions of normality and variance were assessed using standard diagnostic approaches. For variables showing evidence of overall site differences (p < 0.05), pairwise between-site post hoc comparisons were conducted with Holm adjustment to control the family-wise error rate. For variables that allowed selection of more than one answer (e.g., motivations for research participation), the proportion of participants who selected each category were reported and compared across sites. Recruitment and enrollment totals were also reported. Findings were interpreted descriptively given the exploratory study design. All analyses were conducted in R (version 4.4.3).

## RESULTS

### Recruitment and characteristics of recruited population

Data were collected on 12 dates over an 11-month period. Of 187 participants screened, 118 were eligible, and 85 enrolled (Figure 3). Enrolled participants were primarily retired, highly educated, non-Hispanic, White females with moderate OA (Table 2). The sample contained similar proportions of participants with bilateral and unilateral OA. A broad spectrum of income ranges and education levels was observed at the Senior Center, although no statistical differences in income or other demographic characteristics were observed across sites. Forty percent of the sample included participants who were either new to research and/or enrolled as walk-ins.

**Figure 3.**
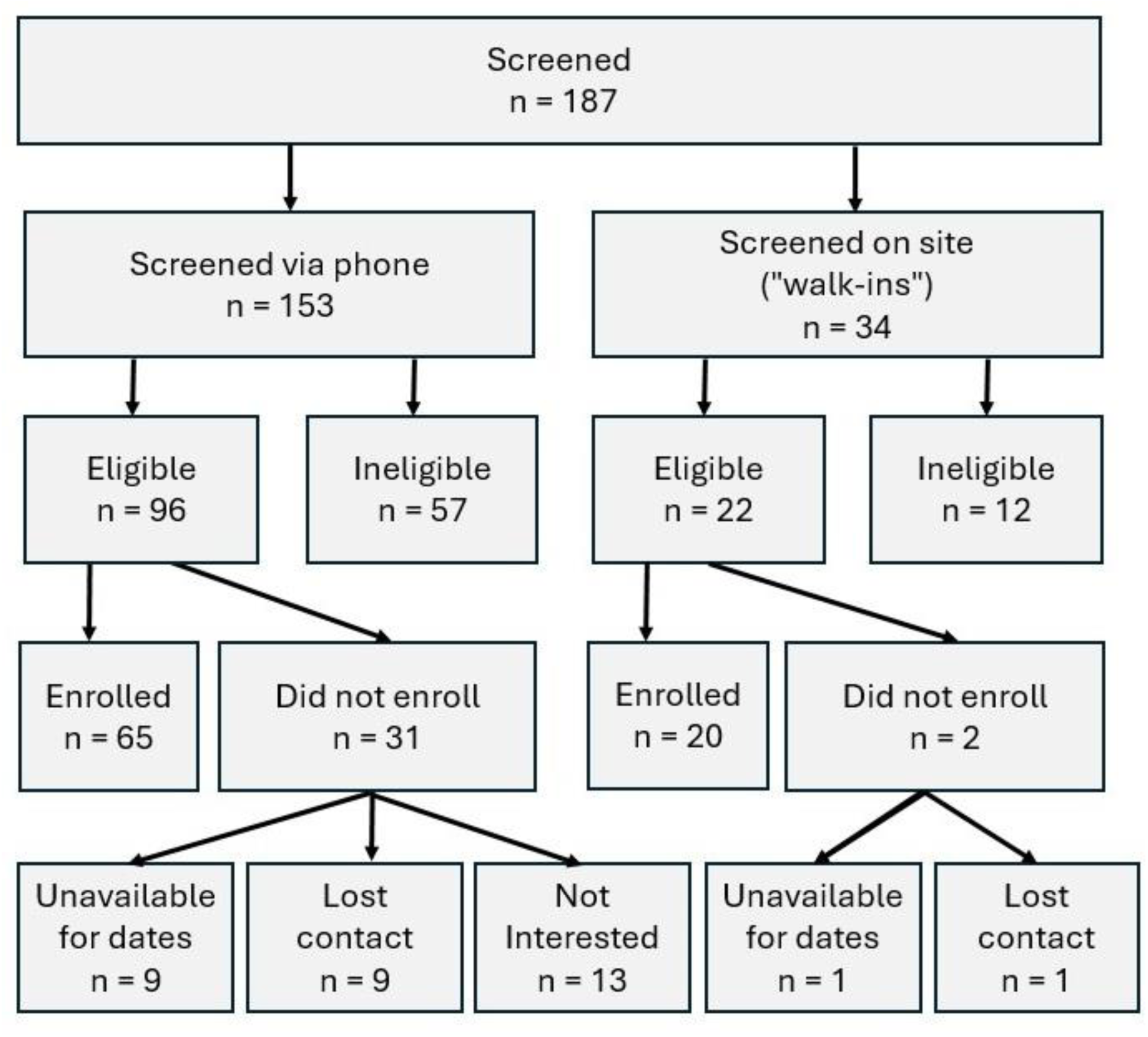
Study flow diagram.

**Table 2.**
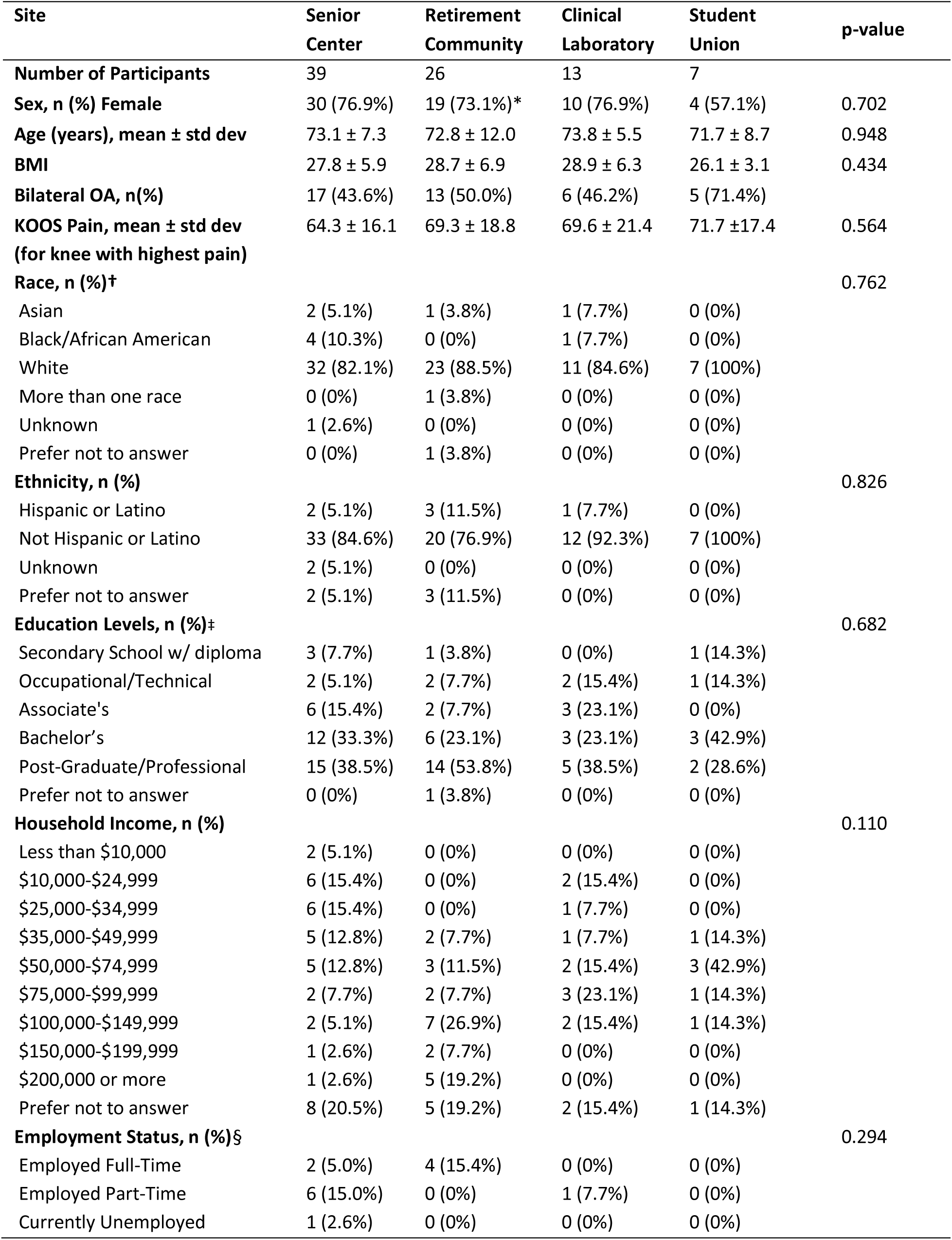
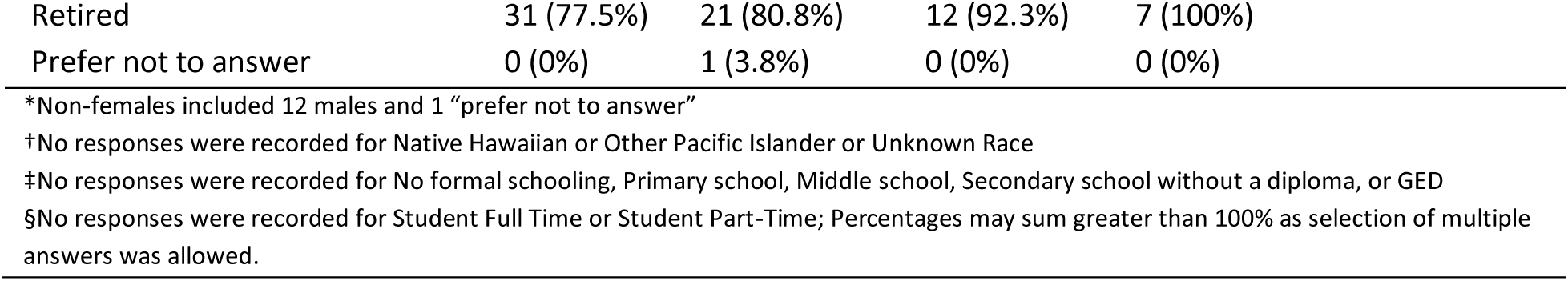
Participant characteristics by site.

| Site | Senior Center | Retirement Community | Clinical Laboratory | Student Union | p-value |
| --- | --- | --- | --- | --- | --- |
| **Number of Participants** | 39 | 26 | 13 | 7 |  |
| **Sex, n (%) Female** | 30 (76.9%) | 19 (73.1%)* | 10 (76.9%) | 4 (57.1%) | 0.702 |
| **Age (years), mean ± std dev** | 73.1 ± 7.3 | 72.8 ± 12.0 | 73.8 ± 5.5 | 71.7 ± 8.7 | 0.948 |
| **BMI** | 27.8 ± 5.9 | 28.7 ± 6.9 | 28.9 ± 6.3 | 26.1 ± 3.1 | 0.434 |
| **Bilateral OA, n(%)** | 17 (43.6%) | 13 (50.0%) | 6 (46.2%) | 5 (71.4%) |  |
| **KOOS Pain, mean ± std dev (for knee with highest pain)** | 64.3 ± 16.1 | 69.3 ± 18.8 | 69.6 ± 21.4 | 71.7 ± 17.4 | 0.564 |
| **Race, n (%)†** |  |  |  |  | 0.762 |
| Asian | 2 (5.1%) | 1 (3.8%) | 1 (7.7%) | 0 (0%) |  |
| Black/African American | 4 (10.3%) | 0 (0%) | 1 (7.7%) | 0 (0%) |  |
| White | 32 (82.1%) | 23 (88.5%) | 11 (84.6%) | 7 (100%) |  |
| More than one race | 0 (0%) | 1 (3.8%) | 0 (0%) | 0 (0%) |  |
| Unknown | 1 (2.6%) | 0 (0%) | 0 (0%) | 0 (0%) |  |
| Prefer not to answer | 0 (0%) | 1 (3.8%) | 0 (0%) | 0 (0%) |  |
| **Ethnicity, n (%)** |  |  |  |  | 0.826 |
| Hispanic or Latino | 2 (5.1%) | 3 (11.5%) | 1 (7.7%) | 0 (0%) |  |
| Not Hispanic or Latino | 33 (84.6%) | 20 (76.9%) | 12 (92.3%) | 7 (100%) |  |
| Unknown | 2 (5.1%) | 0 (0%) | 0 (0%) | 0 (0%) |  |
| Prefer not to answer | 2 (5.1%) | 3 (11.5%) | 0 (0%) | 0 (0%) |  |
| **Education Levels, n (%)‡** |  |  |  |  | 0.682 |
| Secondary School w/ diploma | 3 (7.7%) | 1 (3.8%) | 0 (0%) | 1 (14.3%) |  |
| Occupational/Technical | 2 (5.1%) | 2 (7.7%) | 2 (15.4%) | 1 (14.3%) |  |
| Associate's | 6 (15.4%) | 2 (7.7%) | 3 (23.1%) | 0 (0%) |  |
| Bachelor's | 12 (33.3%) | 6 (23.1%) | 3 (23.1%) | 3 (42.9%) |  |
| Post-Graduate/Professional | 15 (38.5%) | 14 (53.8%) | 5 (38.5%) | 2 (28.6%) |  |
| Prefer not to answer | 0 (0%) | 1 (3.8%) | 0 (0%) | 0 (0%) |  |
| **Household Income, n (%)** |  |  |  |  | 0.110 |
| Less than \$10,000 | 2 (5.1%) | 0 (0%) | 0 (0%) | 0 (0%) | |
| \$10,000-\$24,999 | 6 (15.4%) | 0 (0%) | 2 (15.4%) | 0 (0%) | |
| \$25,000-\$34,999 | 6 (15.4%) | 0 (0%) | 1 (7.7%) | 0 (0%) | |
| \$35,000-\$49,999 | 5 (12.8%) | 2 (7.7%) | 1 (7.7%) | 1 (14.3%) | |
| \$50,000-\$74,999 | 5 (12.8%) | 3 (11.5%) | 2 (15.4%) | 3 (42.9%) | |
| \$75,000-\$99,999 | 2 (7.7%) | 2 (7.7%) | 3 (23.1%) | 1 (14.3%) | |
| \$100,000-\$149,999 | 2 (5.1%) | 7 (26.9%) | 2 (15.4%) | 1 (14.3%) | |
| \$150,000-\$199,999 | 1 (2.6%) | 2 (7.7%) | 0 (0%) | 0 (0%) | |
| \$200,000 or more | 1 (2.6%) | 5 (19.2%) | 0 (0%) | 0 (0%) | |
| Prefer not to answer | 8 (20.5%) | 5 (19.2%) | 2 (15.4%) | 1 (14.3%) |  |
| **Employment Status, n (%)§** |  |  |  |  | 0.294 |
| Employed Full-Time | 2 (5.0%) | 4 (15.4%) | 0 (0%) | 0 (0%) |  |
| Employed Part-Time | 6 (15.0%) | 0 (0%) | 1 (7.7%) | 0 (0%) |  |
| Currently Unemployed | 1 (2.6%) | 0 (0%) | 0 (0%) | 0 (0%) |  |

|  |  |  |  |  |
| --- | --- | --- | --- | --- |
| Retired | 31 (77.5%) | 21 (80.8%) | 12 (92.3%) | 7 (100%) |
| Prefer not to answer | 0 (0%) | 1 (3.8%) | 0 (0%) | 0 (0%) |

\*Non-females included 12 males and 1 “prefer not to answer”
†No responses were recorded for Native Hawaiian or Other Pacific Islander or Unknown Race
‡No responses were recorded for No formal schooling, Primary school, Middle school, Secondary school without a diploma, or GED
§No responses were recorded for Student Full Time or Student Part-Time; Percentages may sum greater than 100% as selection of multiple answers was allowed.

At the community sites, 34 participants walked in and were screened on-site. Of these, 19 enrolled and participated at the same location on the same day. One was screened on-site at a community location, declined participation at the time, and later elected to participate on campus. The percentage of eligible participants who enrolled was higher for those screened on-site as compared to those screened over-the-phone (91% and 68%, respectively). Of 21 individuals reporting no prior research experience, 16 participated at a community site. Notably, 11 of the 20 participants who ultimately enrolled via walk-in pathways were new research participants. Most participants (65%) reported travel times under 30 minutes. Alternative forms of transportation were more commonly reported at community sites, but this was not statistically different across sites (Table 3).

**Table 3.**
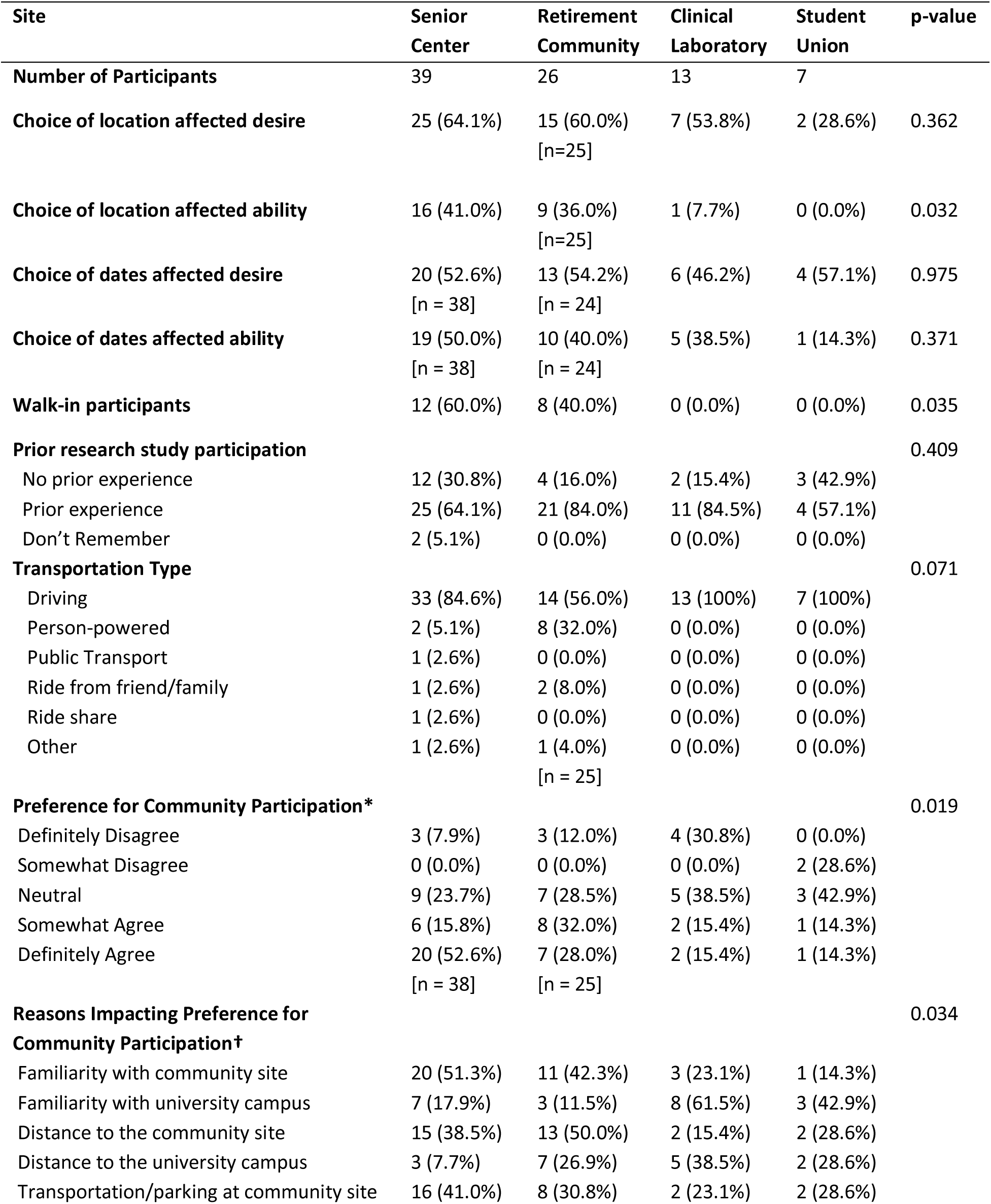
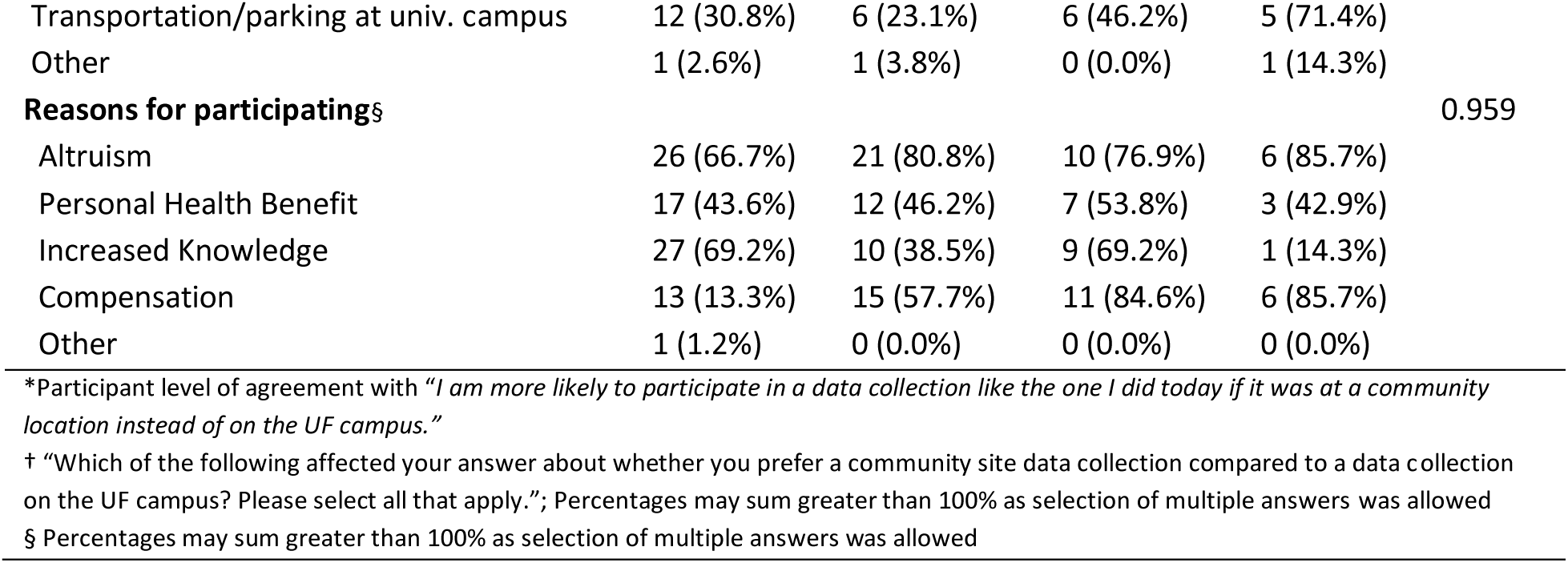
Participant responses to acceptability measures from the custom acceptability questionnaire. Values are reported as cases (% of the group who reported yes). When the number of responses is not equal to the number of participants, sample size is noted below the item.

### Acceptability of community-based data collection

Responses to the question: “*Rate your level of agreement with this statement: I am more likely to participate in a data collection like the one I did today if it was at a community location instead of on the UF campus“* and the reasons selected as affecting this response differed across sites (Table 3). Post hoc comparisons, although not significant, indicate that this effect may come from potential differences between the Senior Center and the Clinical Laboratory (adjusted p = 0.214) or the Student Union (adjusted p = 0.136). Participants primarily indicated familiarity with the site as a reason for their response, but parking/transport options and distance to the site of participation were also important considerations.

While not different across sites, approximately 50% of participants reported that having a choice of date or location affected their desire to participate (Table 3). There was a significant difference across sites in whether a choice of location affected ability to participate (p = 0.032). No other differences in acceptability measures were observed across sites. Although pairwise post-hoc comparisons were not statistically significant, the overall tests indicate heterogeneity in variables across sites.

## DISCUSSION

This study demonstrates the feasibility of a high-throughput community-based biomechanics data collection protocol in individuals with knee OA, with findings indicating that this approach is acceptable and may support broader participation in biomechanics research. Contrary to prior work suggesting that community-based research approaches may increase demographic diversity in research participation,^14, 15^ we did not observe statistically significant differences in demographic characteristics across sites. The recruited population is reflective of the area in which this study was conducted^20^ (where all sites were in relatively close proximity to a major university; Figure 1), with a high proportion reporting travel times of 30 minutes or less. While our sample reflects key features of the broader knee OA population, including a predominance of older adults and women,^21^ this finding may indicate that more purposeful sampling through the identification of community sites that represent the diversity in this geographic region is needed to reduce selection bias and increase representativeness of the sample.^5^ Notably, implementation of the community-based protocol substantially expanded participation through walk-in enrollment and inclusion of new research participants, which together comprised 40% of the enrolled sample of 85 participants. The high number of walk-ins enrolled at community sites (91% of eligible walk-ins; Figure 3) and high proportion of first-time research participants support the accessibility of this data collection model and suggests that community-based implementation may reduce barriers to initial research engagement. Conducting data collection outside the lab does not fully eliminate selection bias,^5^ but the ability to recruit a relatively large sample that is broadly representative of the surrounding community supports further exploration of community-based biomechanics assessment methods.

Overall, participants at the Senior Center—the site with the highest number of walk-ins and new research participants—tended to report greater agreement that they would be more likely to participate in this type of data collection at a community location than on campus (Table 3). Participant responses at the other three sites clustered around ‘neutral’. Familiarity with the data collection location and distance and transportation and parking options emerged as the two most-commonly cited factors influencing this preference (Table 3). Thus, individuals with knee OA may be more likely to participate in research at a familiar, accessible location. This finding is consistent with prior work suggesting that transportation is a common barrier to clinical research participation in older adults^22^ and that community-based research may increase engagement by reducing such barriers.^13^

Consistent with these preferences, participants at the community sites more frequently reported that having a choice of location affected their ability to participate, whereas those attending on-campus sites largely disagreed (Table 3). Overall, 58% of the population reported that a choice of location affected their desire to participate, and this was more pronounced among participants at the community sites. This pattern likely reflects the greater visibility of the community sites and the high number of walk-in participants, as well as more frequent use of alternative transportation methods (i.e., person-powered, public transport, ride-share, golf cart) at these locations. Although scheduling constraints have been cited as a common barrier to research participation in other populations^23^, we found only 41% of the population reported that having a choice of date affected their ability to participate, with lower rates of agreement at the on-campus sites; however, 52% of the population reported that having a choice of date did affect their desire to participate, with no significant differences across sites. Together, these results indicate that this population may be more likely to participate in a research study when given a choice of location and date, and challenges with availability may not play a critical role when offered these choices.

The higher number of scheduled dates and enrolled participants at the community sites may reflect participant site preference, although no formal statistical test of site preference was conducted. Anecdotally, participants often mentioned previous connections to a specific site (e.g., prior clinic visits at the building containing the Clinical Laboratory, regular attendance at the Senior Center, or residing at the Retirement Community), which may have influenced site selection. Notably, the distinction between “community” and “on campus” locations may not have been uniformly perceived by participants. For example, the Clinical Laboratory site was interpreted by some individuals as community-based due to its location on the outer edge of campus. These contextual factors highlight the complexity of defining and interpreting “community-friendly” participation within heterogenous environments.

While altruism appeared to be a common reason for research participation across sites, a high proportion of participants at the on-campus sites indicated that compensation was equally as important. In contrast, participants at the Senior Center reported that an increase in health knowledge was a strong motivation for participation, suggesting that sharing results with the public may be beneficial when working towards “community-friendly” research approaches.

It should also be noted that participant experiences varied across data collection dates and sites. Higher-volume dates were characterized by a more active, social environment and occasional wait times, while lower-volume dates offered a quieter, more individualized experience. Late/early arrivals, walk-ins, and variability in data collection segment duration influenced wait times (described elsewhere) and may have affected the participant experience. Since the order of data collection segments was not fixed, participant impressions may also have been influenced by whether biomechanics or questionnaires were completed first. Last, dates selected for data collections were based on both research team and site availability.

Consequently, some interested individuals may have been unable to participate (Figure 2), including those affected by illness, conflicting appointments, travel, or other commitments. Ultimately, implementation of a high-throughput data collection model introduces natural day-to-day variation that may shape participant experiences.

Overall, these findings suggest that a high-throughput, community-based approach to biomechanics research can increase sample size and enhance research engagement among individuals with knee OA by offering additional recruitment strategies and broadening participation options. Familiarity with, and proximity to, the data collection site appear to be important considerations for this population. Publicly accessible community locations with substantial foot traffic may further encourage participation from individuals who might not otherwise engage in biomechanics research. This approach supports more inclusive and scalable investigations of OA biomechanics, which may help counter bias and homogeneity issues inherent in traditional lab-based research.

## Data Availability

De-identified data may be made available from the corresponding author upon reasonable request, subject to institutional approvals and data use agreements, in accordance with IRB protocols and participant consent.

## ACKNOWLEDGMENTS

We thank the study participants for contributing their time and the staff at the community sites for allowing use of their spaces and assisting with room scheduling and on-site logistics.

We acknowledge the contributions of over 25 AI Biomechanics Laboratory students who assisted with study piloting, logistics, and data collection, in particular graduate students Alexandra Chertok, Alexander Gruber, Peter Schaefer, and Kaitlin Southern. We also thank Kaitlyn Downer and Collin Larke from collaborating laboratories for assisting with data collection during summer sessions.

We acknowledge Clinical Research Core support from the Claude D. Pepper Older Americans Independence Center at the University of Florida, including early guidance on study design and regulatory preparation from Dr. Stephen Anton, study coordination (including recruitment outreach, participant screening, and reminder calls) from Maryam Sohooli, and early coordination support from Nevena Stanojevic.

We acknowledge the insightful comments and discussion provided by Dr. Staja Booker and Dr. Jennifer Nichols during the final stages of manuscript preparation.

## AUTHOR CONTRIBUTIONS

KEC conceived and designed the study, oversaw all aspects of data collection and analysis, obtained funding, and takes responsibility for the integrity of the work as a whole. JMQ led the analyses, coordinated and participated in data collection, contributed to data interpretation, and drafted the initial manuscript. RCM contributed to study design, coordinated and participated in data collection, contributed to analysis and interpretation, and assisted with manuscript drafting. KAS contributed to the development of acceptability measures, interpretation of findings, and critical revision of the manuscript for important intellectual content. PQ and ZT provided statistical expertise, contributed to the analysis and interpretation of data, and critically revised the manuscript for important intellectual content. HKV contributed to study design, interpretation of results, and critical revision of the manuscript. All authors approved the final version for submission.

## DECLARATION OF GENERATIVE AI IN SCIENTIFIC WRITING

During the preparation of this work the authors used ChatGPT (GPT-5.3; OpenAI) during final manuscript editing in order to improve language clarity and flow. After using this tool, the authors reviewed and edited the content as needed and take full responsibility for the content of the publication.

